# Intermittent theta burst stimulation modulates working memory-related theta-gamma coupling in adolescents with ADHD

**DOI:** 10.64898/2026.07.13.26357958

**Authors:** Brian C. Kavanaugh, Megan M. Vigne, Christopher Legere, Zachary Borden, Elena Lynott, Daniel Cheong, Alexa Warren, W. Luke Acuff, Eric Tirrell, Elena K. Festa, Stephanie R. Jones, Richard N. Jones, Anthony Spirito, Linda L. Carpenter

## Abstract

**Objective:** Working memory (WM) deficits are a co-occurring feature to numerous neuropsychiatric disorders, particularly attention-deficit/hyperactivity disorder (ADHD), and there remain no treatments that directly target WM. The coupling between the phase of theta band activity and amplitude of gamma band activity (i.e., TGC) is an established neural correlate of WM. However, no studies have examined WM-related TGC in ADHD or whether neuromodulation can modulate these oscillatory dynamics in youth. This set of studies examined the effects of intermittent theta burst stimulation (iTBS) to the left dorsolateral prefrontal cortex (DLPFC) and left posterior parietal cortex (PPC) on TGC in youth with ADHD.

**Methods:** In two randomized, double-blind, sham-controlled crossover trials, adolescents with ADHD and clinically significant parent-reported WM symptoms first completed a single-session study comparing DLPFC versus PPC iTBS targeting (n = 47) and then a multi-session clinical trial comparing 10 sessions of active versus sham left DLPFC iTBS (n = 29). Participants completed a computerized visuospatial Sternberg WM task with concurrent electroencephalography (EEG) before and after the single sessions, as well as at baseline, midway through treatment, and approximately 24 hours after the final session within the multi-session trial. Phase-amplitude coupling between theta phase and gamma amplitude was quantified using the Kullback–Leibler modulation index at frontoparietal electrodes. Linear mixed-effects models examined treatment effects and associations between change in TGC and WM status (including accuracy, reaction time, and clinical symptoms).

**Results:** Across participants, lower TGC was associated with lower symptoms and better WM performance, including higher accuracy, faster and more consistent RT. Active iTBS increased frontoparietal TGC relative to sham stimulation, with effects observed both acutely after a single session and ∼24 hours after multiple sessions. DLPFC-targeted iTBS increased TGC, whereas PPC-iTBS had no measurable effect. Change in TGC was associated with change in WM, such that a decrease in TGC was associated with faster RT and decreased RT variability. Higher baseline TGC was associated with greater improvement in WM. Active iTBS decoupled the TGC-WM association observed during sham iTBS, and greater electric field intensity of iTBS was associated with greater improvement in WM accuracy and greater decrease in TGC.

**Conclusions:** Active iTBS to the left DLPFC modulated WM-related TGC in youth with ADHD. These findings provide preliminary evidence that neuromodulation may improve WW by modifying oscillatory dynamics within frontoparietal networks. Larger clinical trials with higher stimulation doses are needed to determine whether targeting oscillatory coupling represents a potential therapeutic strategy for WM deficits.

**Trial Registrations:** NCT05102864; NCT05662280

## INTRODUCTION

ADHD is the most frequently diagnosed childhood mental disorder and less than 10% of people with ADHD will experience full recovery by adulthood ^1–3^. Neurocognitive deficits are a well-established feature of ADHD, with small to large effect size deficits observed across intellectual, academic, memory, speed, attention, and executive functioning (EF) domains ^4–6^. While no single neurocognitive deficit can explain all cases of ADHD, attention and EF consistently show the largest effect sizes relative to controls ^4,5^. When aggregating meta-analytic findings across cognitive domains, WM specifically demonstrates the largest effect sizes differentiating individuals with ADHD from controls ^4^. Further, stimulant medications such as methylphenidate have the smallest effect on WM compared to other EF domains ^7^. ADHD is associated with delayed cortical maturation, particularly in prefrontal regions ^8^. WM is known to be subserved by the frontoparietal network (FPN) and its posterior parietal and dorsolateral prefrontal cortical hubs (PPC and DLPFC, respectively), with the PPC responsible for spatial/sensory encoding and the DLPFC responsible for executing cognitive control demands ^9–11^. Prior neuroimaging WM studies have shown increased activation of the FPN during WM demands ^12, 13, 14^ and the degree of FPN activation relates to WM task performance ^14,15^.

Neural oscillations are rhythmic patterns of brain activity measurable with electroencephalography (EEG) that reflect coordinated neuronal firing^16^. “Cross-frequency coupling” is a principled foundation of neuronal communication and brain organization, and specifically refers to when multiple frequencies are present, the phase of slow oscillations modulates the power of fast oscillations ^17^. Theta phase and gamma amplitude coupling (TGC) is a naturally occurring phase-amplitude coupling that is an established neural marker of WM, conceptualized as stimuli being held ‘in mind’ via fast gamma cycles within a slower theta cycle ^18,19^. Using scalp EEG in humans, higher WM-related TGC is known to be a) associated with greater WM load ^20^ and better accuracy ^21^ in healthy adults, and b) the TGC-WM accuracy relationship is longitudinally stable in adulthood ^22^. Abnormalities in TGC have been observed across multiple neuropsychiatric conditions. For example, schizophrenia/psychosis is associated with excessive resting-state TGC ^23,24^ yet insufficient TGC during WM demands ^21^. Alternatively, resting-state TGC in schizophrenia/psychosis is found to be lower than in healthy controls^25,26^. As no studies have examined TGC during WM demands in ADHD, it remains unknown whether excessive or insufficient TGC during WM demands underlies WM-related deficits in ADHD. This is particularly relevant given the known developmental changes in oscillatory dynamics that occur in childhood ^27^.

Transcranial magnetic stimulation (TMS) is a non-invasive neuromodulation technique that uses electromagnetic induction to modulate cortical excitability ^28,29^. When magnetic pulses are applied in a “repetitive” manner (rTMS) daily for multiple weeks, rTMS is an established evidence-based clinical treatment for multiple adult neuropsychiatric disorders and is known to be safe in youth ^30–35^. Intermittent theta burst stimulation (iTBS) is a brief burst-pattern rTMS protocol that delivers a standard dose in three minutes and has been shown to be non-inferior to the standard 10 Hz rTMS protocol for depression ^36^. Multiple meta-analytic findings have shown a small to medium effect sized improvement in WM after rTMS to the DLPFC in healthy, neurological, and neuropsychiatric adult samples ^37–40^. Long-term, sustained positive effects on WM were found to be stronger than acute effects, and the left DLPFC was found to be superior to the right DLPFC for WM improvement ^41^. Further, prior work has shown that iTBS is superior to 10 Hz rTMS in improving WM ^42^.

Oscillatory dynamics reflect one potential mechanism by which iTBS exerts its effects ^43^. TGC has been specifically linked to iTBS as the iTBS parameters of 5 Hz intervals (i.e., every 200 ms) and 50 Hz pulses is thought to mimic TGC-related oscillatory dynamics ^44,45^. There is some evidence that personalizing iTBS to person-specific TGC can lead to better WM outcomes ^46^, and one study found that iTBS to the temporal cortex increased resting-state TGC ^47^. Further, one 10-Hz rTMS study found that one week of daily DLPFC-rTMS in MCI reduced TGC during WM demands, and that lower TGC was associated with higher WM accuracy^48^. Together, TGC reflects a promising target to engage via neuromodulation modalities to remediate WM deficits.

Despite the promise of iTBS for modulating WM-related brain circuits, no studies to date have examined whether rTMS or iTBS can modulate WM-related oscillatory coupling in youth with ADHD. We therefore conducted two randomized, sham-controlled, double-blind, crossover trials investigating the effects of iTBS on TGC in adolescents with ADHD. One single-session study compared DLPFC versus PPC iTBS targeting, and a multi-session clinical trial compared active/sham DLPFC-targeted iTBS on TGC. We hypothesized that active iTBS to the DLPFC would modulate TGC relative sham iTBS and PPC-iTBS, and that the magnitude of TGC modulation would be associated with changes in WM.

## METHODS

### Design

Data here are from two separate double-blind, sham-controlled crossover design clinical trials investigating iTBS for WM in adolescent ADHD at our institution. Current results reflect the primary EEG aim analyses of both studies. In the single-session study (called “TEMP”; NCT05662280; Figure **1.A**) participants received one session of active iTBS (and one session of sham iTBS on a separate day; one-week washout in-between sessions) to either the left DLPFC or the left PPC. Participants completed EEG during a WM task immediately before and after each iTBS session (**Figure 2.A**). In the multi-session clinical trial (called “TB2”; NCT05102864; **Figure 1.B**), participants received 10 once-daily sessions of active iTBS to the left DLPFC in one phase. Participants also completed a phase of 10 once-daily sessions of sham iTBS, with six-week or longer washout in between phases (participants were randomized to active or sham iTBS in phase 1 [and subsequently sham or active in phase 2]). EEG/WM analyzed here were collected before (“pre-iTBS”), halfway (“halfway”), and ∼24 hours after each iTBS 10-session phase (“post-iTBS”; **Figure 2.B**). Local Institutional Review Board approval and an investigational device exemption (IDE) from the United States Food and Drug Administration (FDA) were obtained separately for each trial. We recently showed the clinical symptom efficacy of TB2 ^49^.

**Figure 1.**
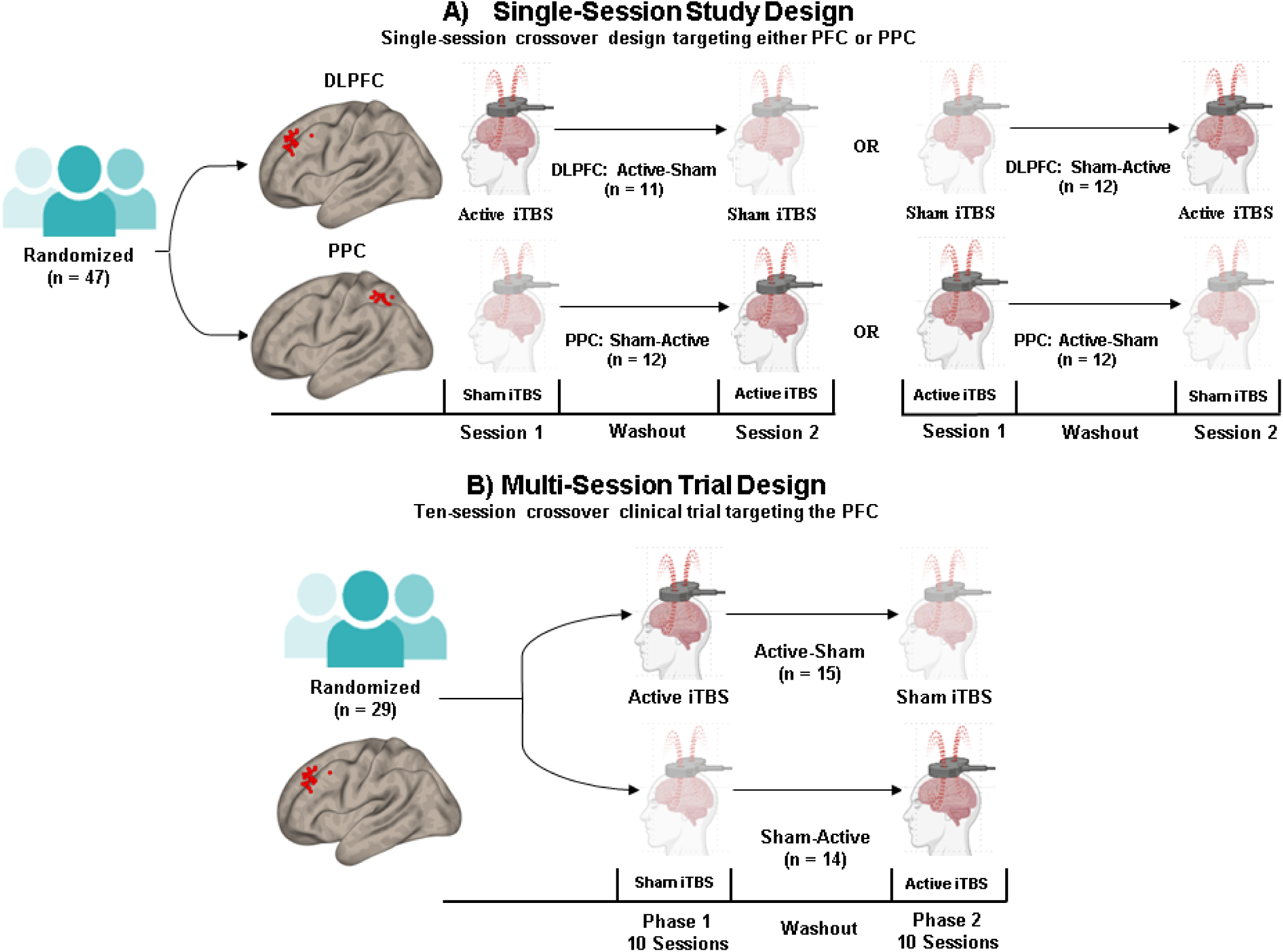
**Legend.** Study design layout of the two described clinical trials investigating theta-burst stimulation for adolescent working memory deficits. Study A (NCT05102864) was a single-session study of left dorsolateral prefrontal cortex (DLPFC) versus left posterior parietal cortex-targeted iTBS (PPC; sham-controlled). Study B (NCT05662280) was a ten-session clinical trial of left DLPFC-targeted iTBS (sham-controlled). Both studies were randomized, double-blind, sham-controlled and crossover design trials. This figure was produced with select images from BioRender.

**Figure 2.**
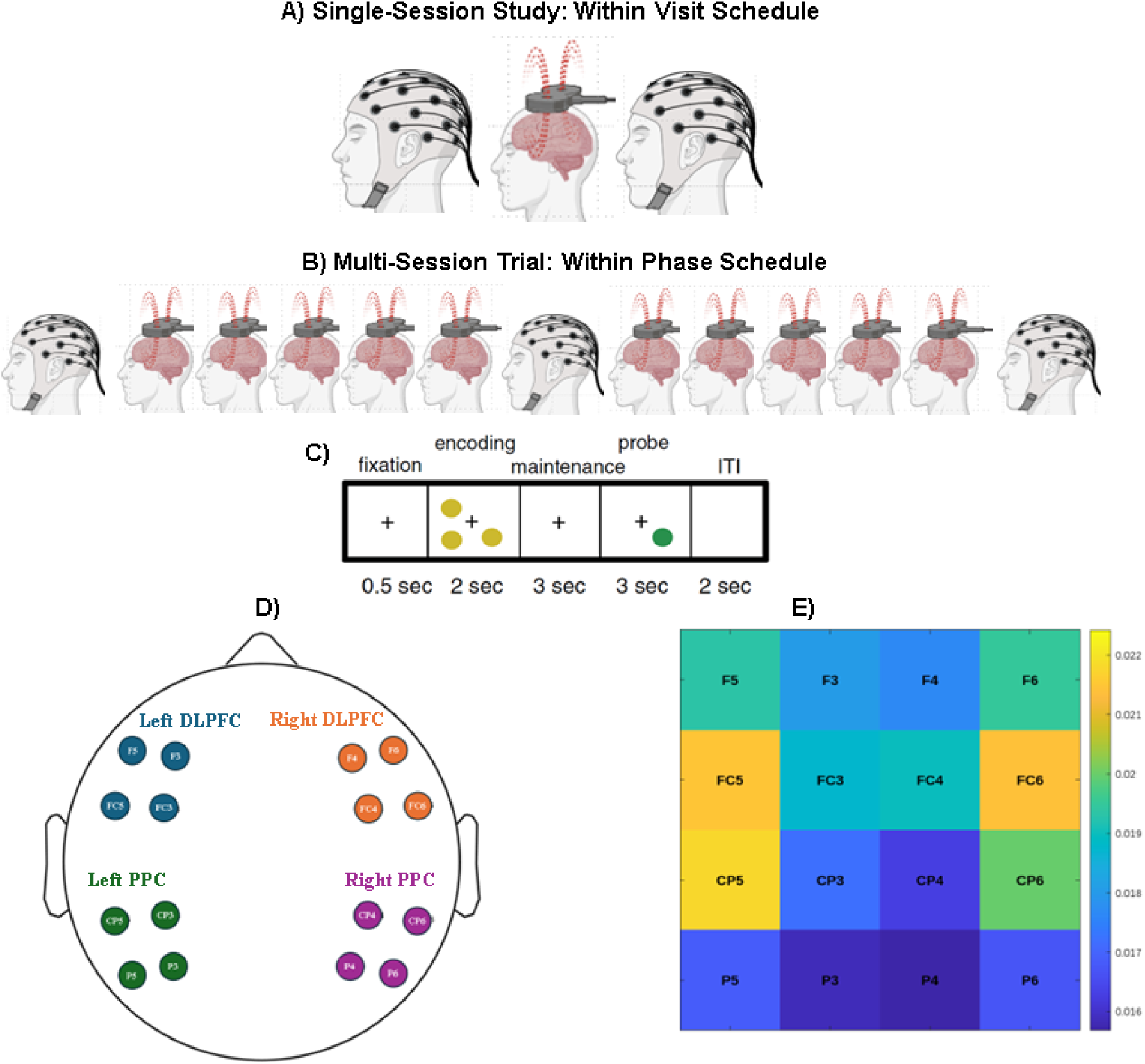
**Legend.** A) Within visit EEG/iTBS schedule for the single-session study. B) Within phase EEG/iTBS schedule for the multi-session trial. C) The experimental paradigm in both studies. Yellow dots were used as encoding stimuli, with one green dot as the stimuli during the probe. As previously described by Lenartowicz et al. (2014). D) The scalp montage of analyzed electrodes across left DLPFC, right DLPFC, left PPC, and right PPC. E) Matrix reflecting the mean TGC metric during WM maintenance across electrodes. This figure was produced with select images from BioRender.

### Participant Sample

Primary <u>inclusion criteria</u> for both studies were: a) a clinical diagnosis of ADHD (confirmed with parent-reported symptoms on the Vanderbilt Assessment Scales), b) parent-reported WM symptoms (i.e., t-score≥ 60 on the Behavior Rating Inventory of Executive Function–Second Edition [BRIEF-2]), c) full-scale intelligence quotient ≥ 80 (based on Wechsler Abbreviated Scales of Intelligence, Second Edition [WASI-II]), and English fluency. <u>Exclusion criteria</u> included active mania, psychosis, substance use, current suicidal ideation, and standard TMS and MRI contraindications including seizure history. Participants continued clinical treatments, including psychostimulants, but were asked to maintain medication consistency throughout the trial. Whether the participant was taking a psychostimulant as usual during EEG/WM procedures was coded as ‘stimulant status’: yes/no.

For TEMP, fifty adolescents (12-18 years old) with ADHD were enrolled and 47 were randomized into the protocol from November 2021 to July 2025 (Table 1). Two participants were determined to be ineligible after enrollment and prior to randomization, two participants withdrew prior to receiving iTBS (including one that was randomized but was unable to tolerate motor threshold calculation), and two participants withdrew after receiving one session of iTBS. There were a total of 6 adverse events (consisting of mild headaches, scalp discomfort, shoulder pain; no serious AEs) for a per-person AE rate of 13%. Forty-four participants completed all study procedures (94% completion rate) and analyses were conducted using the full intent-to-treat sample. For TB2, thirty-four adolescents (12-18 years old) with ADHD were enrolled and 29 were randomized into the protocol from November 2021 to October 2024 (**Table 1**). Twenty-four participants completed the full trial (n = 5 withdrew from the trial; 82% completion rate) and analyses were conducted using the full intent-to-treat sample. As noted elsewhere, there were a total of 11 adverse events (no serious AEs) for a per-person AE rate of 20%. Thirty-three of the thirty-four adolescents that enrolled in TB2 also enrolled in TEMP. TEMP iTBS sessions were completed prior to TB2 iTBS sessions (separated by at least one week).

**Table 1.**
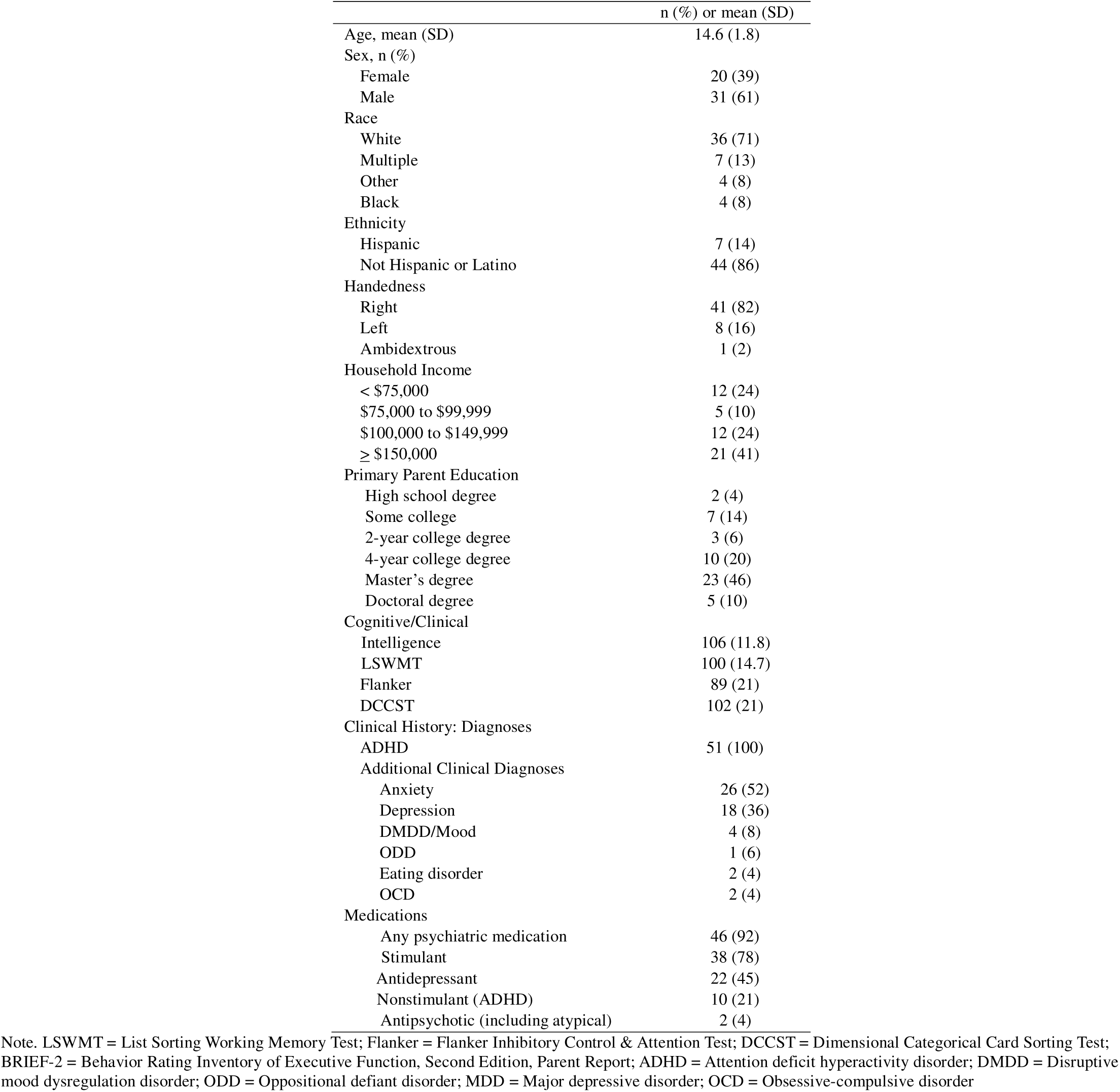
Demographic, clinical, and treatment characteristics of total TEMP-TB2 combined sample (n = 51)

|  | n (%) or mean (SD) |
| --- | --- |
| Age, mean (SD) | 14.6 (1.8) |
| Sex, n (%) |  |
| Female | 20 (39) |
| Male | 31 (61) |
| Race |  |
| White | 36 (71) |
| Multiple | 7 (13) |
| Other | 4 (8) |
| Black | 4 (8) |
| Ethnicity |  |
| Hispanic | 7 (14) |
| Not Hispanic or Latino | 44 (86) |
| Handedness |  |
| Right | 41 (82) |
| Left | 8 (16) |
| Ambidextrous | 1 (2) |
| Household Income |  |
| < \$75,000 | 12 (24) |
| \$75,000 to \$99,999 | 5 (10) |
| \$100,000 to \$149,999 | 12 (24) |
| ≥ \$150,000 | 21 (41) |
| Primary Parent Education |  |
| High school degree | 2 (4) |
| Some college | 7 (14) |
| 2-year college degree | 3 (6) |
| 4-year college degree | 10 (20) |
| Master's degree | 23 (46) |
| Doctoral degree | 5 (10) |
| Cognitive/Clinical |  |
| Intelligence | 106 (11.8) |
| LSWMT | 100 (14.7) |
| Flanker | 89 (21) |
| DCCST | 102 (21) |
| Clinical History: Diagnoses |  |
| ADHD | 51 (100) |
| Additional Clinical Diagnoses |  |
| Anxiety | 26 (52) |
| Depression | 18 (36) |
| DMDD/Mood | 4 (8) |
| ODD | 1 (6) |
| Eating disorder | 2 (4) |
| OCD | 2 (4) |
| Medications |  |
| Any psychiatric medication | 46 (92) |
| Stimulant | 38 (78) |
| Antidepressant | 22 (45) |
| Nonstimulant (ADHD) | 10 (21) |
| Antipsychotic (including atypical) | 2 (4) |

Note. LSWMT = List Sorting Working Memory Test; Flanker = Flanker Inhibitory Control & Attention Test; DCCST = Dimensional Categorical Card Sorting Test; BRIEF-2 = Behavior Rating Inventory of Executive Function, Second Edition, Parent Report; ADHD = Attention deficit hyperactivity disorder; DMDD = Disruptive mood dysregulation disorder; ODD = Oppositional defiant disorder; MDD = Major depressive disorder; OCD = Obsessive-compulsive disorder

### Treatment Intervention

For all iTBS sessions, stimulation was delivered using a Nexstim NBT System 2 with a figure-eight coil. The iTBS protocol consisted of 2-second trains of triplet 50-Hz bursts repeated every 10 seconds (600 pulses/session) at 80% resting motor threshold, with sessions lasting approximately three minutes. Sham stimulation produced <2% of the active coil electric field. Stimulation targeted the left DLPFC (TEMP & TB2) or PPC (TEMP) using individual MRI-guided neuronavigation. The left DLPFC was targeted based on anatomical landmark method described by Mylius et al ^50^. The left PPC was targeted based on Kiriyama et al ^51^, specifically the horseshoe shaped angular gyrus (AG) of the left inferior parietal lobule (IPL; **Figure 1**). In TB2, induced electric field estimates (V/m; generated by the Nexstim software) were averaged across sessions (not calculated for TEMP as it was only a single session).

### Working Memory Task

All participants completed a computerized, visuospatial Sternberg WM paradigm (**Figure 2.C**) ^52,53^. Each trial included a 0.5-second fixation, a two-second encoding period with 1, 3, 5, or 7 yellow dots, a three-second delay, and a probe period during which a single green dot was presented for up to three seconds. Participants indicated whether the probe matched any prior dot location by pressing “same” or “different.” The task was administered using E-Prime 3.0 and included practice trials. In TEMP, as the task was completed immediately before and after a single iTBS session, participants completed a 10-minute version of the task comprising 72 trials across two blocks with equal load distribution of only 3, 5, or 7 yellow dot stimuli. In TB2, participants completed the standard 14-minute task comprising 96 trials across two blocks with equal load distributions across 1, 3, 5, or 7 yellow dot stimuli. Raw accuracy and mean reaction time, and mean reaction time standard deviation were utilized, consistent with prior work with this task ^53^. Load conditions were collapsed for analyses. All data points were included.

### WM Symptoms

In TB2, parent-reported clinical WM symptoms were gathered at the same timepoints as EEG/WM. WM symptoms were assessed with the Behavior Rating Inventory of Executive Functions – Second Edition (BRIEF-2), Working Memory Subscale (i.e., eight items, rated never, sometimes, often) ^54^.

### EEG Recording and Pre-Processing

EEG recording was obtained during the Sternberg WM task with a 64-electrode montage integrated in a neoprene cap (Waveguard by ANT Neuro). Recording was conducted at 2048 Hz sampling rate with CPZ set as reference electrode using full-band EEG DC amplifier. Pre-processing was conducted using the EEGLAB toolbox for MATLAB. Data was resampled to 250 Hz, line noise was removed (using the *Zapline* plugin in EEGLAB), then high-pass filtered at 1 Hz and low-pass filtered at 80 Hz using zero-phase filters. EEGLAB’s *clean_rawdata* function was used to remove channels when a) signal was flat for 5+ seconds, b) high-frequency line noise was >4 standard deviations (SDs) above the mean, and c) when there was < .80 correlation with nearby channels. This function also performed artifact subspace reconstruction with a burst rejection threshold of 20 SD outside the mean and removed bad data periods with > 25% of channels out of acceptable range. Rejected channels were spherically interpolated and data was re-referenced to average. Independent component analysis (ICA) was conducted with the picard algorithm. Using EEGLAB’s *ICLabel*, components with ≥ .8 probability of eye or muscle artifact were rejected. An epoch was rejected (using EEGLAB’s pop_autorej) if it contained a value outside the ± 100uV range (starting probability = 5, maximum = 5). Data were epoched relative to the fixation onset preceding each trial, with a baseline correction of -.5 to 0 (reflecting the 500 ms prior to fixation) to capture fixation (500 milliseconds), encoding (2000 milliseconds), and the maintenance or “delay” (3000 milliseconds).

### Theta-Gamma Coupling

Phase-amplitude coupling was calculated within the theta (4-7 Hz) and gamma (30-50 Hz) bands via the PACTools extension in EEGLAB ^55^. PACTools calculates the extent to which the phase of a low-frequency oscillation modulates the amplitude of a high frequency oscillation. This study utilized the Kullback–Leibler Modulation Index (KLMI) ^56^. TGC was calculated in frontoparietal regions, specifically the left DLPFC (F3, F5, FC3, FC5), right DLPFC (F4, F6, FC4, FC6), left PPC (P3, CP3, P5, CP5), and right PPC (P4, CP4, P6, CP6; **Figure 2.E**). This electrode selection was based on the known frontoparietal involvement of WM-related dynamics ^9–11^. TGC was calculated for the fixation, encoding, and maintenance stages of each trial. Phase frequencies were sampled using four logarithmically spaced bins between 4 and 7 Hz, and amplitude frequencies were sampled using seven logarithmically spaced bins between 30 and 50 Hz. KLMI values were averaged across frequency bins to generate one TGC value per epoch, and extracted for the mean of fixation, encoding, and maintenance phases. TGC values were averaged within each region of interest and then across frontoparietal regions (DLPFC-PPC TGC; **Figure 2.F**).

TGC is known to be highly size-dependent ^20,56^. To address the concern that restricting analyses to only correct trial epochs would lead to an over-inflated correlation with overall task accuracy, all available epochs (including correct and incorrect trials) were included in analyses. Follow-up analyses examining correct/incorrect responses or different WM load were not conducted due to size dependence concerns. Further, select prior work has calculated TGC within ten random trials of available data instead of all available data due to concerns about size-dependence ^20^. We calculated TGC using both ‘all trials’ and ’10 random trials’ approaches here. In TB2, the ‘all trials’ and ’10 random trials’ TGC calculations were highly correlated to one another (r = .74, p < .001) and displayed a nearly identical association to WM accuracy (r = -.57 & =-.59, both p < .001; Fisher’s z test = .25, p = .40). In TEMP, the ‘all trials’ and ’10 random trials’ TGC calculations were again highly correlated to one another (r = .76, p < .001) and displayed a similar association to WM accuracy (r(143) = -.34 & -.19, p < .001 & p = .027, respectively; Fisher’s z test = -1.4, p = .09). We therefore utilized the ‘all trials’ approach to maximize validity with all available data.

### Data Availability

In TEMP, EEG/WM data was available for n = 41 participants for a total of n = 150 participant sessions (including n = 77 for visit 1 [37/40 for pre/post iTBS] and n = 73 for visit 2 [37/36 for pre/post iTBS]), out of the n = 46 and n = 180 administered participant/sessions (83% of administered EEG/WM procedures available for analysis). Two of the n = 150 (1.3%) of the datapoints were removed as these files had < 20 trials (4 & 12) and resulted in TGC metrics > 3 SD from the sample mean. In TB2, EEG/WM data was available for n = 26 participants for a total of n = 140 participant sessions (phase 1 pre-iTBS = 25, phase 1 halfway = 24, phase 1 post-iTBS = 24, phase 2 pre-iTBS = 24, phase 2 halfway = 21, phase 2 post-iTBS = 22, out of the n = 29 and n = 156 administered participant sessions (90% of administered EEG/WM procedures available for analysis). Two outliers (1% of data) were detected for TGC z-scores and winsorized to z = 3.0.

### Statistical Analyses

Primary linear mixed-models examined TGC. To characterize TGC patterns, models included fixed effects of stage and region, while controlling for treatment (active vs. sham), visit, and pre/post session, with a random intercept for participant. Significant effects were followed by pairwise comparisons. Linear mixed-models examined the effect of iTBS on absolute change in TGC (TEMP: post–pre; TB2: halfway–pre and post–pre), with fixed effects of order (sham-active/active-sham), target (DLPFC/PPC; TEMP only), condition (active/sham), visit (TB2 only), TGC electrode region (DLPFC/PPC), TGC stage (fixation/encoding/maintenance), and their interactions, and a random intercept for participant. Follow-up linear mixed models examined the association between change in TGC and WM, with fixed effects including condition, target (TEMP only), baseline WM, baseline TGC, TGC change, and their interactions. Specific to WM symptoms, age and sex were included as covariates given the known role of these variables in WM symptom expression, and only post-iTBS (not halfway) was analyzed as change in symptom expression was not expected to be as acutely observed as direct WM performance change. Pearson correlation analyses examined the association between average e-field intensity per person (controlling for motor threshold) and change in TGC/WM. Analyses were conducted in SPSS. Statistical significance was set at p < .05.

## RESULTS

### Theta-Gamma Coupling

A linear mixed-effects model (controlling for active/sham iTBS, visit, and pre/post session) indicated a significant effect of stage (F[2, 1665] = 16.5, p < .001) and region (F[3, 1746] = 44.6, p < .001), but not a stage x region interaction on TGC (F(6, 1746) = 0.35, p = .913). Follow-up pairwise comparisons indicated that there was no difference in TGC during fixation and encoding stages, but TGC during the maintenance stage was lower than fixation and encoding. Further, TGC at the DLPFC electrodes was higher than TGC at the PPC electrodes (**Figure 3.A**). Across participants (in TEMP given larger sample size), lower average TGC (maintenance) was associated with older age (r = -.43, p = .007), better WM accuracy (r = -.41, p = .01), faster reaction time (RT; r = .36, p = .03), and reduced RT variability (RTSD or RT variability; r = .44, p = .005; **Figure 3.A**). Across participants (in TB2 given repeated symptom assessment), lower average TGC was associated with lower WM symptoms (r = .44, p = .026; **Figure 3.A**).

**Figure 3.**
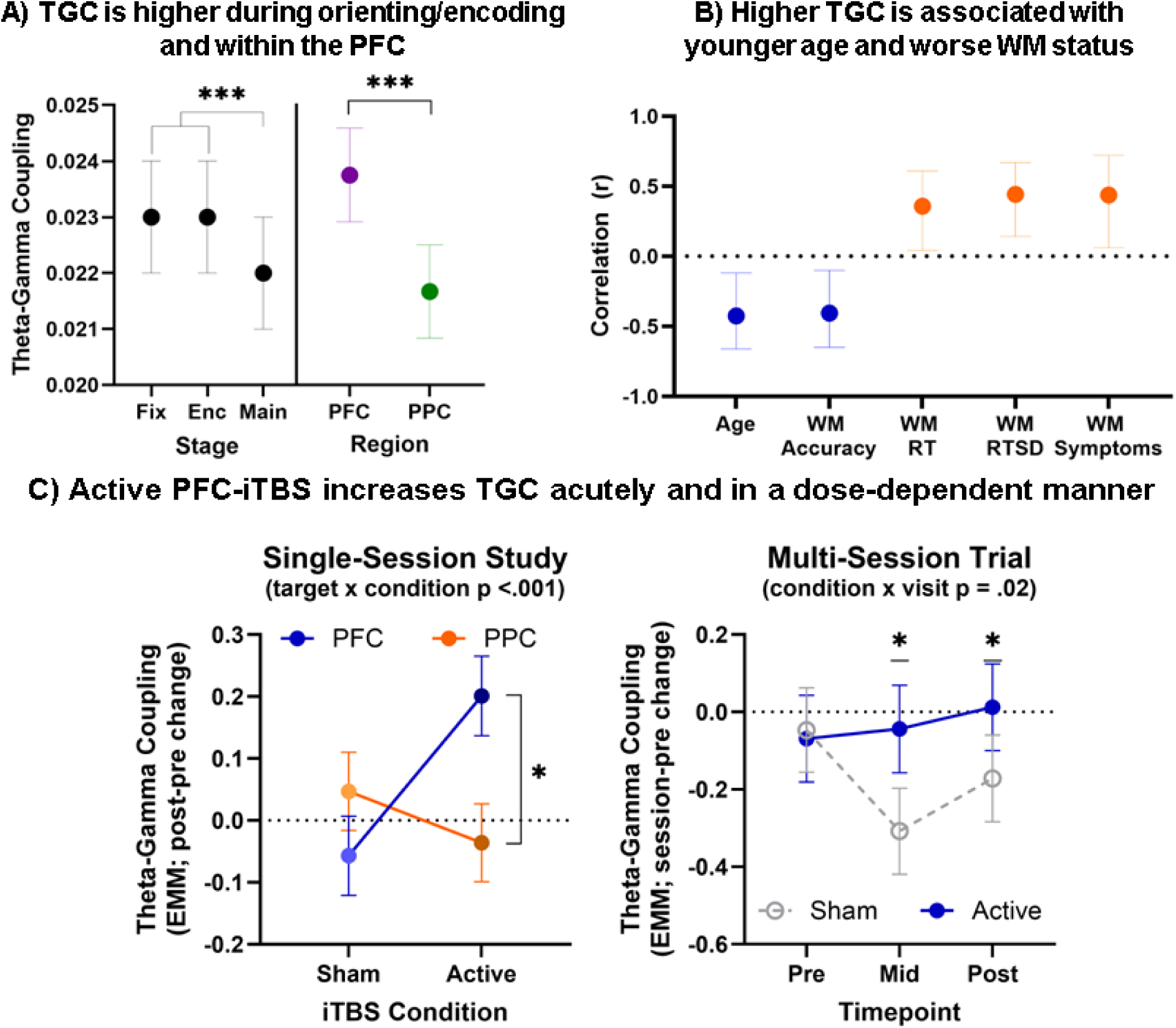
**Legend.** A) A mixed linear model with stage, region, and their interaction as fixed effects showed that TGC was lower during the maintenance stage and higher in the DLPFC region (no interaction). B) A series of person-mean correlation analyses showed that higher TGC was associated with younger age, lower WM task accuracy, higher WM task reaction time (RT), higher WM task RT standard deviation, and higher parent-reported WM symptoms. C) Primary mixed linear models showed that active iTBS to the left DLPFC increased TGC in both acute and sub-acute patterns.

### Single-Session Study (TEMP): Primary Effects of iTBS

Linear mixed-effects models examined the effect of iTBS on absolute change in TGC (i.e., post – pre iTBS score). There was a significant effect of condition (F[1,381] = 5.2, p = .02) and a significant condition x target interaction (F[1,381] = 20, p < .001; **Table 2.A**). No primary effect or interaction with TGC electrodes or TGC stage indicates that the effect of iTBS on TGC was stable across regions and stages. Follow-up pairwise comparisons indicate that active iTBS was superior to sham iTBS in increasing TGC, and that active iTBS to the DLPFC was superior to both sham-PFC and active-PPC in increasing TGC (**Figure 3.C**, **left**).

**Table 2.** Linear mixed model effects on theta-gamma coupling and working memory.

| A) TEMP: TGC (Post-Pre Change) |  |  |  |
| --- | --- | --- | --- |
|  | df | F | p |
| Intercept | 1,35 | .90 | .35 |
| Order (sham-active/active-sham) | 1,35 | 1.3 | .26 |
| iTBS Target (DLPFC/PPC) | 1,34 | .67 | .42 |
| **Condition (active/sham)** | **1,381** | **5.2** | **.02** |
| TGC Electrodes (DLPFC/PPC) | 1,370 | .01 | .94 |
| TGC Stage (Fixation/Encoding/Maintenance) | 2, 370 | 1.2 | .32 |
| **Condition x Target** | **1, 381** | **20** | **<.001** |
| Condition x Target x Stage | 6, 370 | .35 | .91 |
| Condition x Target x Electrodes | 3, 370 | .63 | .60 |
| B) TB2: TGC (Session-Pre Change) |  |  |  |
|  | df | F | p |
| Intercept | 1,25 | .42 | .53 |
| Order (sham-active/active-sham) | 1,25 | .04 | .84 |
| **Condition (active/sham)** | **1,781** | **10.1** | **.002** |
| Visit | 2,776 | 2.7 | .07 |
| TGC Electrodes (DLPFC/PPC) | 1,774 | .80 | .37 |
| TGC Stage (Fixation/Encoding/Maintenance) | 2, 774 | .43 | .65 |
| **Condition x Visit** | **2, 777** | **3.9** | **.02** |
| Condition x Visit x TGC: DLPFC-PPC | 5, 774 | 1.2 | .33 |
| Condition x Visit x TGC: Stage | 10, 774 | .24 | .99 |

Note. Tx = treatment. DLPFC = Dorsolateral prefrontal cortex. PPC = Posterior parietal cortex. Session reflects halfway time point and post-iTBS time point.

**Table 3.** TEMP: Working memory – TGC associations (post-pre change)

|  | Accuracy |  |  | Reaction Time (RT) |  |  | RT Standard Deviation |  |  |
| --- | --- | --- | --- | --- | --- | --- | --- | --- | --- |
|  | df | F | p | df | F | p | df | F | p |
| Intercept | 1,26 | .08 | .78 | 1,59 | 11.2 | .001 | 1,33 | 3.0 | .09 |
| Condition | 1,26 | .51 | .48 | 1,59 | .43 | .52 | 1,30 | 2.0 | .17 |
| Target | 1,22 | .53 | .48 | 1,59 | 3.3 | .07 | 3,31 | .59 | .45 |
| Baseline WM | 1,24 | 4.6 | .037 | 1,59 | 8.4 | .005 | 1,58 | 17.6 | <.001 |
| Baseline TGC | 1,55 | 1.3 | .25 | 1,59 | 4.0 | **.049** | **1,59** | **5.0** | **.03** |
| TGC Change | 1,58 | 1.7 | .20 | **1,59** | **5.2** | **.026** | 1,58 | .01 | .94 |
| Condition x TGC Change | 1,53 | 1.0 | .32 | 1,59 | .23 | .63 | 1,59 | .17 | .68 |
| Target x TGC Change | 1,39 | .00 | .98 | 1,59 | .01 | .92 | 1,52 | .002 | .97 |
| Condition x Target x TGC Change | 1,52 | .61 | .44 | 1,59 | .38 | .54 | 1,58 | 1.3 | .25 |

Note. WM = Working memory. TGC = Theta gamma coupling.

**Table 4.**
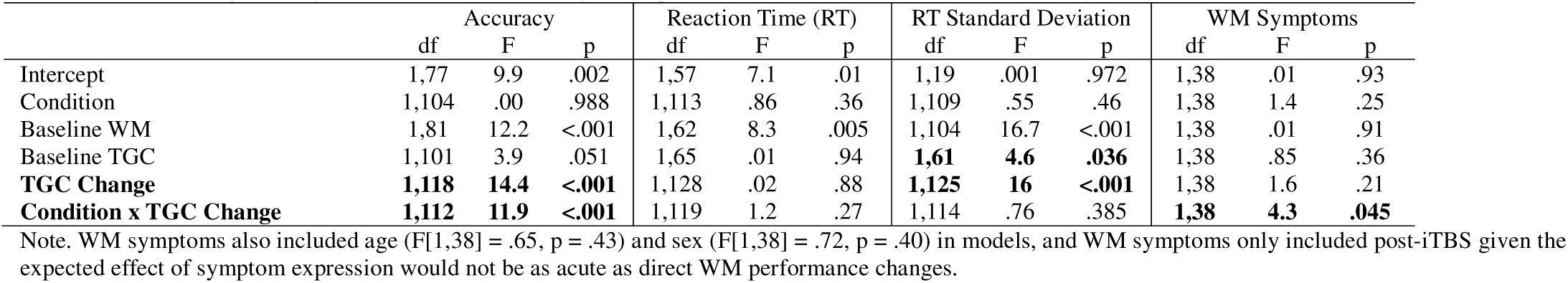
TB2: Working memory – TGC associations (change from pre-iTBS)

### Multi-Session Clinical Trial (TB2): Primary Effects of iTBS

Linear mixed-effects models examined the effect of iTBS on absolute change in TGC (i.e., session – pre iTBS score). There was a significant effect of condition (F[1,781] = 10.1, p = .002) and a significant condition x visit interaction (F[2,777] = 3.9, p = .02; **Table 2.B**). No primary effect or interaction with TGC electrodes or TGC stage indicates that the effect of iTBS on TGC was stable across regions and stages. Follow-up pairwise comparisons indicate that active iTBS was superior to sham iTBS in increasing TGC at halfway and post-iTBS timepoints (**Figure 3.C, right**).

### Association between TGC and WM performance

Linear mixed models examined the association between change in TGC and change in WM across both studies (**Tables 3 & 4**). <u>Change in TGC</u> was significantly associated with change in a) RT (F[1,59] = 5.2, p = .026; TEMP) and b) RTSD (F[1,125] = 16, p < .001; TB2; **Figure 4.A**), in that a decrease in TGC was associated with faster RT/RTSD. <u>Baseline TGC</u> was significantly associated with a) RT (F[1,59 = 4.0, p = .049), b) RTSD in TEMP (F[1,59] = 5.0, p = .03), and c) RTSD in TB2 (F[1,61] = 4.6, p = .036), in that higher baseline TGC was associated with greater improvement in WM. A condition x TGC change interaction was found for a) accuracy (F[1,112] = 11.9, p < .001) and b) WM symptoms (F[1,38] = 4.3, p = .045; **Figure 4.B**), in that active iTBS negated the TGC-WM association found during sham iTBS.

**Figure 4.**
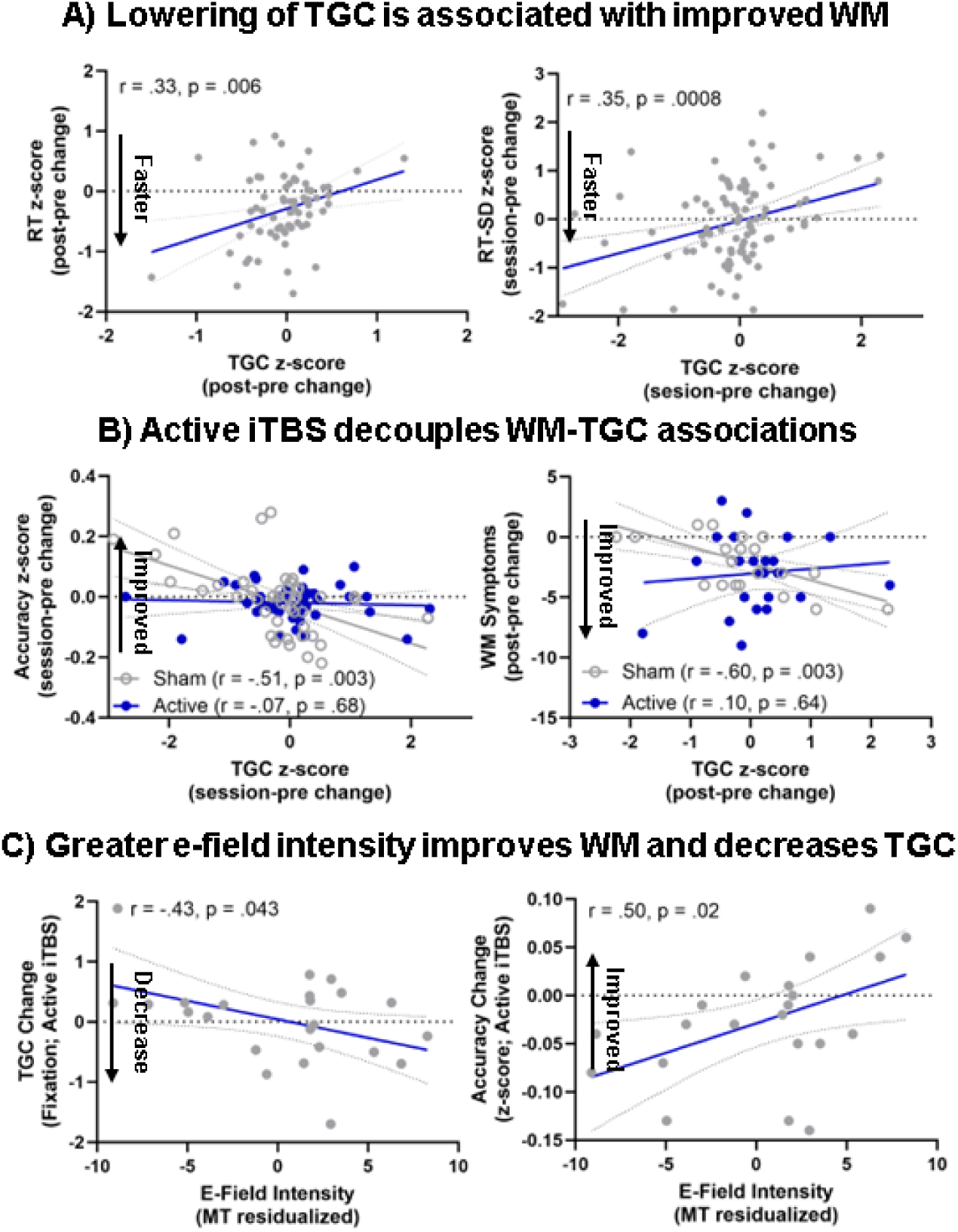
**Legend.** Correlation analyses (follow-up from primary mixed linear model) examining association between TGC change and RT/RTSD change in the single-session study (left) and multi-session trial (right), and the mediating role of active/sham iTBS status (C). Correlation analyses of average electric field intensity (V/m) of active iTBS and TGC (left) and accuracy (right) change during active iTBS.

### E-Field correlates of TGC/WM change

Finally, given the repeated dosing of iTBS in TB2, Pearson correlation analyses examined whether individual variability in stimulation intensity predicted treatment response. Greater average e-field intensity per person (after regressing out motor threshold) was associated with greater improvement in WM accuracy (r = .50, p = .02) and greater decrease in TGC (during fixation; r = -.43, p = .043; Figure **4.C**). But not WM RT, RTSD, or TGC during encoding or maintenance (all p > .05).

## DISCUSSION

These two double-blind, sham-controlled, crossover trials examined the effects of iTBS to the left dorsolateral prefrontal cortex (DLPFC) and posterior parietal cortex (PPC) on theta-gamma coupling (TGC) during working memory (WM) demands in adolescents with ADHD. To our knowledge, these are the first studies to investigate whether transcranial magnetic stimulation can modulate WM-related TGC in youth with ADHD. We found evidence that iTBS to the DLPFC, but not PPC, modulated frontoparietal TGC and that the degree of modulation was associated with WM improvements.

In the first analysis to investigate TGC in youth, we found a unique pattern relative to prior adult findings, in that *lower* TGC was associated with older age, better WM performance, and a lower severity of WM-related clinical symptoms. In healthy adults, *higher* WM-related TGC is associated with higher WM load and better WM performance ^21^, although dynamics may differ in clinical and developmental cohorts. In disorders like schizophrenia, patients show excessive TGC during rest ^23,24^ and insufficient TGC during WM demands ^21^. Alternatively, in ADHD, patients are found to have insufficient TGC during rest ^25,26^ (i.e., the opposite pattern of schizophrenia), but there are limited data on TGC during WM demands. Developmental factors may be highly involved, as for example, elevated gamma bursting during WM demands is found in youth and this gamma bursting linearly decreases during development ^27^. Relatedly, we found that older participants had lower TGC. Together, this emphasizes the role of development in the direction of the TGC-WM relationship, although it remains unknown if this finding is specific to ADHD or generalized across all youth. Current preliminary findings suggest that in youth ADHD, higher TGC during WM demands is associated with worse WM status, potentially reflecting excessive coupling that becomes more fine-tuned and efficient in adulthood.

We found that DLPFC-targeted iTBS increased WM-related TGC, observed both acutely after a single iTBS session, and post-acutely (∼24 hours post-iTBS) after multiple iTBS sessions. iTBS has been theoretically linked to TGC as it administers pulses at theta intervals (i.e., every 200 milliseconds) via gamma intensity (i.e., 50 Hz)^44,45^. However, prior work on the neuromodulation of TGC has been highly inconsistent, and only a small number of studies have shown any effect on TGC outcomes ^43^. One prior study in healthy adults found that a single session of iTBS to the temporal cortex acutely *increased* resting-state TGC ^47^, while another study found that a one-week course 10 Hz rTMS for older adults with MCI *decreased* WM-related TGC and that lower TGC was associated with better WM performance^48^. Our group-level results build upon prior inconsistent findings to show that iTBS can acutely and post-acutely increase WM-related TGC in youth with ADHD.

In comparing the acute effects of iTBS to the left DLPFC or left PPC, we found that DLPFC-targeted iTBS acutely increased TGC and PPC-iTBS had no measurable effect on TGC. At the same time, we found no difference in TGC effects when measured at DLPFC versus PPC electrodes, in that observed effects were consistent across DLPFC and PPC electrodes. The DLPFC and PPC are hubs of the frontoparietal network (FPN) known to underlie WM, with the PPC more involved in sensory encoding and DLPFC more involved in cognitive control ^9–11^. Despite this, nearly all rTMS-WM studies have targeted the DLPFC ^37–40^. In the one study of parietal rTMS for WM, Riddle et al found that a single session of rTMS administered at the alpha frequency had a nuanced positive effect on WM performance ^57^, suggesting that rTMS frequency should be considered in parietal-targeted rTMS for WM. Taken together, here we found that DLPFC-iTBS was superior to PPC-iTBS in acutely modulating TGC and WM, suggesting that the DLPFC remains the most promising target for TGC/WM modulation.

While it may sound initially paradoxical that higher TGC was associated with worse WM status and that iTBS increased TGC, the pattern reflects a clear, yet complex, TGC-WM-iTBS relationship. First, higher baseline TGC was associated with greater improvement in reaction time and variability (RT/SD), in that those participants with excessive baseline TGC had better WM improvements. Similarly, a greater degree of TGC lowering was associated with faster and more consistent responding, suggesting that a lowering of (excessive) TGC led to improved WM. Importantly, for both WM accuracy and WM symptoms, active iTBS decoupled the TGC-WM relationship, in that the strong TGC-WM association during sham iTBS was negligible during active iTBS. This indicates that while iTBS increases TGC as expected, in a cohort characterized by excessive TGC, the pathway to response is more complex than simply increasing TGC leads to improved WM. This is not inconsistent with prior literature, in that the only study on the topic similarly found that a lowering of TGC was associated with improved WM^48^. Taken together, a) iTBS increases TGC at the group level, but b) excessive baseline TGC was associated with worse baseline WM but greater WM improvement, c) the degree to which this excessive baseline TGC is reduced predicts WM response, and d) active iTBS disrupts the typical TGC-WM association.

We found preliminary evidence of a dose-response relationship of iTBS on TGC/WM. Specifically, we found a condition-by-visit interaction for TGC, although similar TGC effects were observed after both five and ten iTBS sessions. Further, higher individual electric field intensity was associated with greater improvement in WM accuracy and greater lowering of TGC, indicating that the dose of iTBS is associated with the subsequent response. Given the overall low dose of iTBS in these studies, this dose-response relationship finding suggests that potentially optimizing iTBS placement, orientation, and/or dose could lead to more pronounced WM improvements. The strong TGC-WM change associations described above further indicate that the degree of target engagement is associated with improved WM status.

Of note, we found no specificity of TGC modulation or TGC-WM relationships to a stage of WM (i.e., fixation, encoding, maintenance) or a specific region (i.e., DLPFC or PPC electrodes). While TGC was higher in the DLPFC compared to the PPC, interestingly, TGC was lower during maintenance than during fixation/encoding, which suggests it is not specifically underlying WM demands (i.e., when stimuli are removed and the individual is required to hold information ‘in mind’). While current results are encouraging and further confirm the utility of TGC, the lack of dynamic change or stage/region specificity suggests that more dynamic oscillatory mechanisms beyond TGC may reflect more promising neural targets of iTBS.

This study has multiple limitations. First, we had no healthy control group to compare TGC findings to that of typically developing youth. While we utilized established approaches for DLPFC/PPC targeting, our results may have limited generalizability and precision, respectively, as we did not utilize standardized Montreal Neurological Institute (MNI) coordinates and we did not utilize functional MRI targeting methods. Additionally, this study had a small sample (particularly in the clinical trial) which limits statistical power and generalizability of findings. The clinical trial utilized a low dose of iTBS given safety concerns (i.e., 600 pulses per day for 10 total sessions), so current findings may be an underestimate of iTBS effects that are achieved with accelerated iTBS protocols that are increasingly used in clinical settings. Finally, no embedded measures of performance validity were administered so true task engagement, effort, and/or validity cannot be confidently stated.

Nevertheless, these findings provide preliminary evidence that iTBS to the left DLPFC can acutely and post-acutely modulate WM-related TGC in youths with ADHD. Active iTBS increased frontoparietal TGC and the degree of modulation correlated with WM change, together suggesting that neuromodulation (particularly iTBS) may potentially improve WM via the modulation of underlying TGC mechanisms. Current findings support the need for larger clinical trials with higher stimulation dosing and improved study design. While the clinical significance of such modulation remains to be determined, these results support further investigation using larger samples and higher stimulation doses.

## Declaration of generative AI and AI-assisted technologies in the manuscript preparation process

During the preparation of this work the authors used ChatGPT to improve readability and identify errors the manuscript. After using this tool/service, the authors reviewed and edited the content as needed and take full responsibility for the content of the published article.

## Data Availability

Available upon request

## Acknowledgements

These trials were funded by the National Institutes of Health (K23MH129853, P20GM130452), NARSAD Brain & Behavior Research Foundation and the Carol Peterson Foundation.

